# An Efficient and Accurate Distributed Learning Algorithm for Modeling Multi-Site Zero-Inflated Count Outcomes

**DOI:** 10.1101/2020.12.17.20248194

**Authors:** Mackenzie J. Edmondson, Chongliang Luo, Rui Duan, Mitchell Maltenfort, Zhaoyi Chen, Kenneth Locke, Justine Shults, Jiang Bian, Patrick B. Ryan, Christopher B. Forrest, Yong Chen

## Abstract

Clinical research networks (CRNs), made up of multiple healthcare systems each with patient data from several care sites, are beneficial for studying rare outcomes and increasing generalizability of results. While CRNs encourage sharing aggregate data across healthcare systems, individual systems within CRNs often cannot share patient-level data due to privacy regulations, prohibiting multi-site regression which requires an analyst to access all individual patient data pooled together. Meta-analysis is commonly used to model data stored at multiple institutions within a CRN; while relatively simple to implement, meta-analysis can result in biased estimation, notably in rare-event contexts. We present a communication-efficient, privacy-preserving algorithm for modeling multi-site zero-inflated count outcomes within a CRN. Our method, a one-shot distributed algorithm for performing hurdle regression (ODAH), models zero-inflated count data stored in multiple sites without sharing patient-level data across sites, resulting in estimates closely approximating those that would be obtained in a pooled patient-level data analysis. We evaluate our method through extensive simulations and two realworld data applications using electronic health records (EHRs): examining risk factors associated with pediatric avoidable hospitalization and modeling serious adverse event frequency associated with a colorectal cancer therapy. Relative to existing methods for distributed data analysis, ODAH offers a highly accurate, computationally efficient method for modeling multi-site zero-inflated count data.

## Introduction

The recent advent of “big data” has had significant implications for health care, spawning several advancements in management and analysis of large-scale patient data [1]. Much of this is a result of the widespread adoption of electronic health records (EHRs), patient data collected during routine and emergency clinical visits. Though EHRs are primarily used as a written record of health care delivery, substantial effort has been made in using these data secondarily to generate real-world evidence (RWE), evidence produced as a result of analyzing observational health data outside of clinical trials. RWE quality can be substantially improved from analyzing pooled data, patient records aggregated across health systems. This is especially true in the context of studying rare outcomes, where outcome prevalence at any single institution may not be large enough to result in an analysis with meaningful conclusions. Pooled patient data from several healthcare systems also allows for study of a sample likely to be more representative of the population of interest.

While pooling patient data from several institutions is ideal, doing so is not always possible. Regulations such as the Health Insurance Portability and Accountability Act (HIPAA) in the United States and the General Data Protection Regulation (GDPR) in the European Union often prevent inter-site sharing of patient-level data that is not de-identified [2-3]. Sharing de-identified individual patient data (IPD) can also be dangerous according to several studies demonstrating the susceptibility of these data to re-identification, causing concern among patients [4-6]. Further, significant computational burden associated with storing and analyzing massive datasets makes data centralization less appealing in some settings. As a result of these restrictions and concerns, much interest has been demonstrated recently in distributed clinical research networks (CRNs), multi-site distributed data networks which allow for analyses across institutions without the need for data centralization [7-8]. In a CRN, each individual institution or health system maintains control over its own data, drastically reducing risk of violating patient privacy through avoiding IPD exchange. Examples of CRNs include the National Patient-Centered Clinical Research Network (PCORnet) [9], a CRN with patient data from 348 health systems in the United States, and the Sentinel System, a national CRN for monitoring performance of FDA-regulated medical products [10-11].

To protect patient privacy, CRNs largely invoke methods for synthesizing and analyzing aggregate data, summary measures obtained from individual sites without any information that could reveal patient identity. In comparative effectiveness research performed in CRNs, meta-analysis is frequently used. Meta-analysis, which only requires effect size and variance estimates from each individual site, is easy to implement and widely accepted in medical literature; it is the primary analysis method used in comparative effectiveness studies conducted by the Observational Health Data Sciences and Informatics (OHDSI) collaborative, an international CRN made up of over 100 different databases [12-16].

While suitable for many applications, meta-analysis has been shown to result in biased or imprecise effect estimates in the context of rare events and limited sample sizes [17]. In only sharing site-level point and variance estimates, meta-analysis does not utilize any additional aggregate information that could be obtained from ongoing studies with access to their own patient-level data. Distributed regression methods are an alternative to meta-analysis which leverage access to patient-level data within individual sites, allowing for fitting a regression model distributively across institutions without sharing IPD. While several distributed regression algorithms have been developed, many require several rounds of communication among sites until convergence, resulting in analysis that is both time consuming and computationally expensive [18-19]. More recently, a class of non-iterative distributed algorithms has been proposed by Duan et al. [17, 20]; these methods use a surrogate likelihood approach to generate estimates comparable to those from pooled analysis using IPD only at the lead site, incorporating aggregate information from collaborating sites to better approximate the complete data likelihood [21]. Methods based on the surrogate likelihood approach are one-shot algorithms, requiring only one or two rounds of non-iterative communication among institutions to offer a communication-efficient alternative for performing distributed regression.

To our knowledge, despite the growing collection of methods for analyzing data in CRNs, no distributed regression method for modeling count outcomes currently exists. Count data are abundant in EHRs, administrative claims, and other sources of electronic health data, with examples including length of stay, number of primary care or emergency department visits, and number of laboratory tests administered. To explore associations between count outcomes and a set of clinical covariates, Poisson or Negative Binomial regression is typically used. In practice, medical count data can be zero-inflated, where zero counts are in excess; zeros often make up the majority of observed counts for rare outcomes, far exceeding the number expected in Poisson or Negative Binomial distributions. In several applications, empirical distributions of zero-inflated counts can also feature a small number of observations with relatively large counts. This is a common occurrence in distributions of health care expenditure, for example, which feature a large proportion of patients with no expenses at one end and a smaller proportion of patients with large expenses at the other [22]. In these settings, one can use hurdle regression, which uses two separate processes for modeling zero and non-zero (positive) counts. The first part models whether an observation will have a zero or positive count, commonly through logistic regression, while the second estimates a count for an observation given that the count is positive, typically using zero-truncated Poisson or Negative Binomial regression. Hurdle regression allows one to separately investigate the effect of covariates on the probability of experiencing an outcome and on the expected frequency of an outcome given that it occurs at least once, improving interpretation in settings where these two processes are driven by different parameters.

We propose a novel method, a one-shot distributed algorithm for hurdle regression (ODAH), to distributively model zero-inflated count outcomes stored in multiple institutions. Using the surrogate likelihood approach, our method for modeling count outcomes is an efficient, non-iterative algorithm which requires two rounds of privacy-preserving communication among sites to generate accurate and precise population-level estimates closely approximating those from pooled analysis. We evaluate ODAH through an extensive simulation study before applying our method to two real-world data use cases: analyzing risk factors of pediatric avoidable hospitalization and modeling serious adverse event frequency for colorectal cancer patients.

## Results

### Overview of ODAH method

Our ODAH method approximates and maximizes the pooled, complete data log-likelihood function using patient-level data from only one care site, the *lead* site, incorporating aggregate information from all other (*collaborating*) sites included in the analysis (Figure 1). ODAH requires only two rounds of non-iterative communication among sites. Briefly, the algorithm begins with each collaborating site fitting a two-part, Poisson-Logit hurdle model.

**Figure 1.**
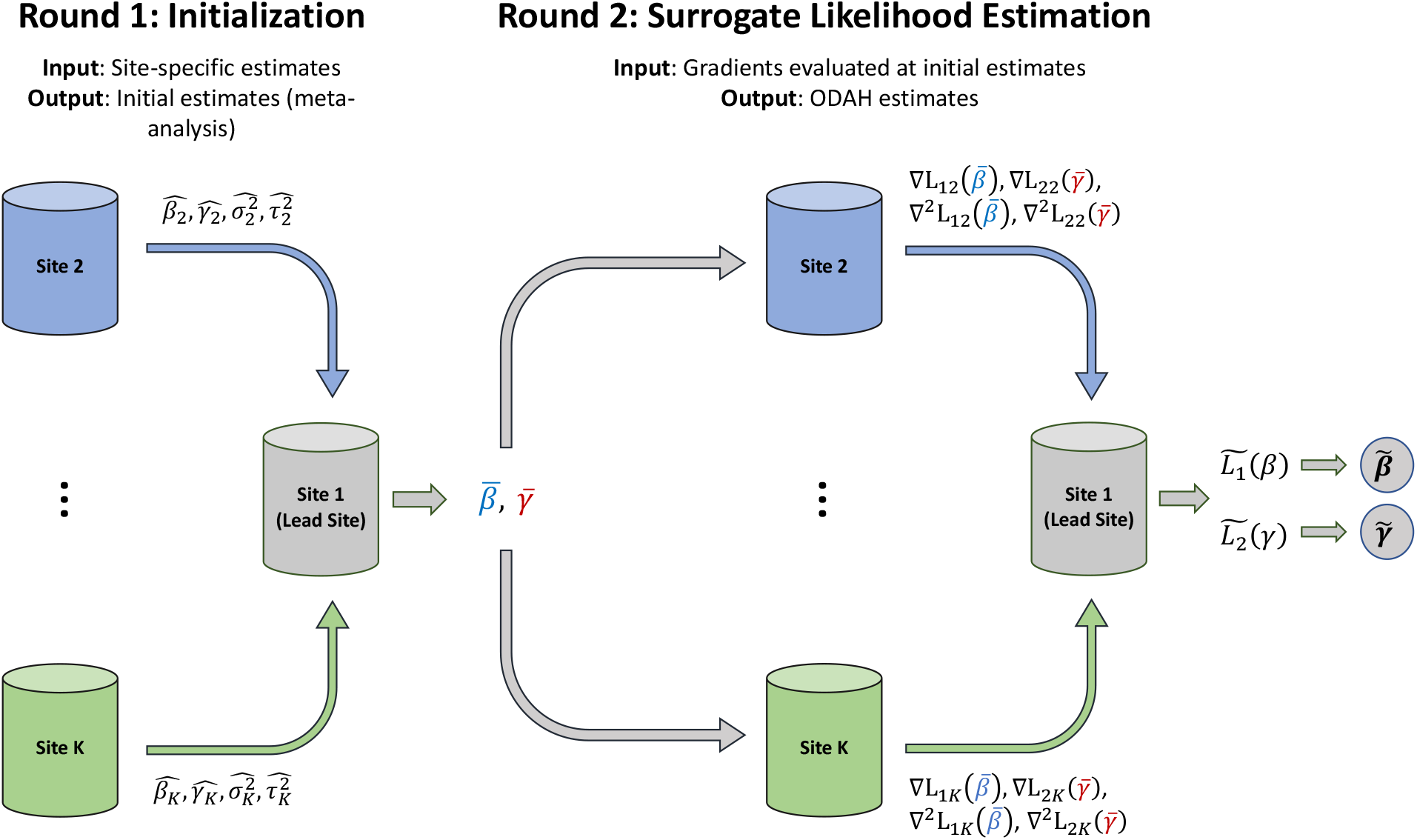
Visual representation of one-shot distributed algorithm for hurdle regression (ODAH). In the initialization round, coefficient 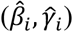 and variance 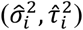 estimates from fitting separate hurdle models at each collaborating site are sent to the lead site; these estimates are then used together with lead site estimates in a meta-analysis to produce initial estimates 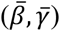 for ODAH, which are sent to each collaborating site. In the surrogate likelihood estimation round, first-order (∇*L*_1*i*_, ∇*L*_2*i*_) and second-order (∇^2^*L*_1*i*_, ∇^2^*L*_2*i*_) gradients are computed at each site, evaluated at the received initial estimates and sent to the lead site. These gradients are used in conjunction with data from the lead site to construct surrogate likelihood functions 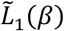 and 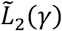, which are then maximized to produce surrogate maximum likelihood estimates 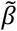 and 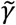.

Coefficient and variance estimates from each component of the hurdle model are sent to the lead site, which in turn synthesizes all individual site estimates (including its own) via meta-analysis to produce initial estimates for ODAH. These initial meta-analysis estimates are then transmitted to the collaborating sites, where first- and second-order gradients evaluated at the initial estimates are calculated for each hurdle model component. These gradients are then sent to the lead site which, through synthesis of lead site patient-level data and collaborating site gradients, formulates and maximizes the surrogate log-likelihood function for each hurdle model component to obtain the ODAH estimates. Full details of the algorithm and derivation of the Poisson-Logit hurdle model are provided in the Methods section.

### Simulation study

To evaluate ODAH empirically in a controlled setting, we conducted a simulation study to primarily compare the performance of ODAH to that of meta-analysis, which does not incorporate any patient-level data. Performance is evaluated in terms of bias relative to pooled estimates, coefficients estimated in an analysis where all patient-level data is used; we treat pooled estimation as our gold standard, an ideal scenario featuring centralized data that is typically unattainable in practice. We additionally examine performance of hurdle regression using data only from the lead site, emulating a single-site analysis.

In our simulations, a count outcome *Y* was associated with two risk factors, *X*_1_ and *X*_2_. *X*_1_ was generated using a truncated Normal distribution emulating the number of primary care visits per year for each patient in our avoidable hospitalization analysis (*X*_1_∼*N*(3, 2), *X*_1_ ∈ (0, 18)), while *X*_2_ was generated using a Bernoulli distribution with the probability of success representing that of public insurance use among patients in the same analysis (*X*_2_∼*Bern*(0.33)). Our covariate of interest was *X*_2_, with *X*_1_ assumed to be a confounder. The outcome *Y* given covariates *X*_1_ and *X*_2_ was generated from the Poisson-Logit hurdle model described in the Methods section, using logistic regression to model the process generating zero or positive counts and zero-truncated Poisson regression to estimate counts given that they are positive. Note that while the hurdle model has two components, each of which can use its own unique set of covariates, the sets of covariates making up each component of the model are identical in our simulations. We seek to estimate ***β*** = {*β*_0_, *β* _1_, *β*_2_} and ***γ*** = {*γ*_0_, *γ* _1_, *γ*_2_}, each 3 x 1 vectors of regression coefficients quantifying associations between our simulated count outcome and risk factors.

Motivated by our rare-event applications, we primarily sought to examine how varying levels of low outcome prevalence and event rate affect the performance of ODAH relative to pooled analysis. We explored four rare-event prevalence settings while holding event rate constant at 0.03 (mean event rate for patients in our avoidable hospitalization analysis, denoting number of hospitalizations per year): 5%, 2.5%, 1%, and 0.5%. To evaluate the effect of event rate on method performance, we explored additional event rates of 0.25, 0.01, and 0.005 while holding outcome prevalence constant at 2.5%. Note that these event rates include zero counts, with smaller event rates corresponding to more severe zero-inflation.

In all settings, we fixed the number of sites *K* = 10 and total population size *N* = 200,000. In settings where we vary outcome prevalence or event rate, we set *n*_1_ = *n*_2_ = ⋯ = *n*_10_ so all sites had the same number of observations. We also explored the effect of the lead site being larger than collaborating sites, setting lead site sizes at 38,000 (collaborating site size 18,000), 56,000 (collaborating site size 16,000), and 74,000 (collaborating site size 14,000). All ten unique simulation settings explored are summarized in Table 1.

**Table 1.** Simulation settings varying baseline outcome prevalence *β*_0_, baseline event rate *γ*_0_, and size of lead site *n*_*lead*_.

| Simulation Setting |  |  | True Parameter Values |  |  |
| --- | --- | --- | --- | --- | --- |
| Prevalence | Event Rate ($\lambda$) | $n_{lead}/N$ | $\beta_0$ | $\gamma_0$ | $n_{lead}$ |
| 5% | 0.03 | 0.10 | -3.0 | -3.6 | 20,000 |
| 2.5% | 0.03 | 0.10 | -3.7 | -3.6 | 20,000 |
| 1% | 0.03 | 0.10 | -4.5 | -3.6 | 20,000 |
| 0.5% | 0.03 | 0.10 | -5.3 | -3.6 | 20,000 |
| 2.5% | 0.25 | 0.10 | -3.7 | -1.4 | 20,000 |
| 2.5% | 0.01 | 0.10 | -3.7 | -4.5 | 20,000 |
| 2.5% | 0.005 | 0.10 | -3.7 | -5.3 | 20,000 |
| 2.5% | 0.03 | 0.19 | -3.7 | -3.6 | 38,000 |
| 2.5% | 0.03 | 0.28 | -3.7 | -3.6 | 56,000 |
| 2.5% | 0.03 | 0.37 | -3.7 | -3.6 | 74,000 |

For each setting, we evaluated estimation accuracy in terms of bias relative to pooled estimates across 1,000 simulations to examine the variability in method performance. In all settings, we assume true coefficient values {*β*_1_, *γ* _1_} = −1 and {*β*_2_, *γ*_2_} = 1.

Figure 2 depicts simulation results from evaluating method performance across all scenarios described in Table 1. Across settings, there was no discernable difference in method performance for estimating *β*_2_, the regression coefficient associated with *X*_2_ in the logistic component of the hurdle model. We therefore present the simulation results for estimating *γ*_2_, leaving *β*_2_ estimation results for the Supplement. Due to select iterations of lead site analysis resulting in outlying estimates, the median bias for the lead site estimate across iterations is reported rather than the mean.

**Figure 2.**
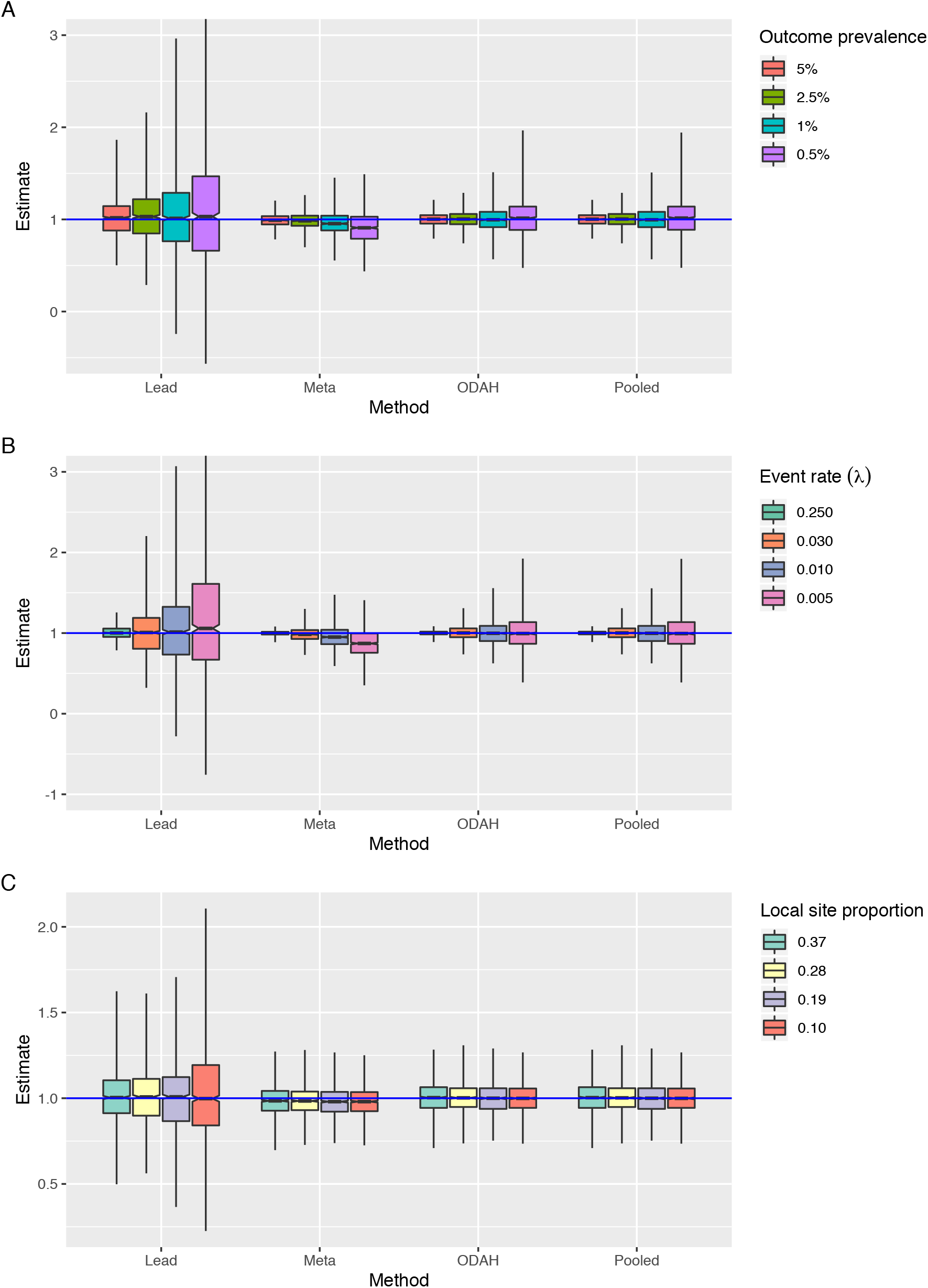
Simulation results for estimating zero-truncated Poisson component covariate *γ*_2_. A) Results for Setting A, fixing *n*_*lead*_ = 20,000 and *γ*_0_ = -3.6 (*λ* = 0.03) while varying outcome prevalence. B) Results for Setting B, fixing *n*_*lead*_ = 20,000 and *β*_0_ = -3.7 (2.5% prevalence) while varying event rate (*λ*). C) Results for Setting C, fixing *β*_0_ = - 3.7 (2.5% prevalence) and *γ*_0_ = −3.6 (*λ* = 0.03) while varying proportion of observations in lead site. Horizontal blue line represents true value of *γ*_2_ = 1.

When lead site size and event rate were fixed at 20,000 and *λ* = 0.03, respectively, we varied outcome prevalence to study how each method performed relative to pooled analysis, the gold standard (Figure 2A). In all prevalence levels examined, ODAH performed nearly as well as pooled analysis, with negligible difference in terms of bias and variance of its estimate; bias in the ODAH estimate relative to the pooled estimate was less than 0.1% for each prevalence level. Conversely, meta-analysis bias relative to the pooled estimate increased with decreasing prevalence, ranging from 0.97% (5% prevalence) to 10.4% (0.5% prevalence). Lead site analysis exhibited the largest variance of all methods; bias relative to the pooled estimate ranged from 0.79% (5% prevalence) to 2.77% (0.5% prevalence).

When lead site size and outcome prevalence were fixed at 20,000 and 2.5%, respectively, we varied event rate to examine its impact on estimating *γ*_2_ in a low prevalence setting (Figure 2B). For all methods, variance of estimates decreased with increasing event rate. ODAH and meta-analysis estimates were nearly identical to pooled estimates when events rates were set to *λ* = 0.25 and 0.03, exhibiting negligible bias relative to the pooled estimate (ODAH bias < 0.1%, meta-analysis bias < 1.9%). When the event rate was set to *λ* = 0.01 and 0.005, ODAH again exhibited negligible relative bias (< 0.1%) but meta-analysis exhibited larger bias relative to the pooled estimate (4.57% and 12.7%, respectively). Lead site analysis exhibited the largest variance of all methods examined, maintaining relatively low relative bias to the pooled estimate when *λ* = 0.25, 0.03 and 0.01 (< 1.1%) but larger bias when *λ* = 0.005 (5.31%).

When examining the effect of increasing lead site size while fixing outcome prevalence and event rate at 2.5% and *λ* = 0.03, respectively, there was not substantial evidence for lead site size affecting ODAH or meta-analysis performance relative to pooled analysis (Figure 3C). Variance of lead site analysis estimates decreased with increasing lead site size.

**Figure 3.**
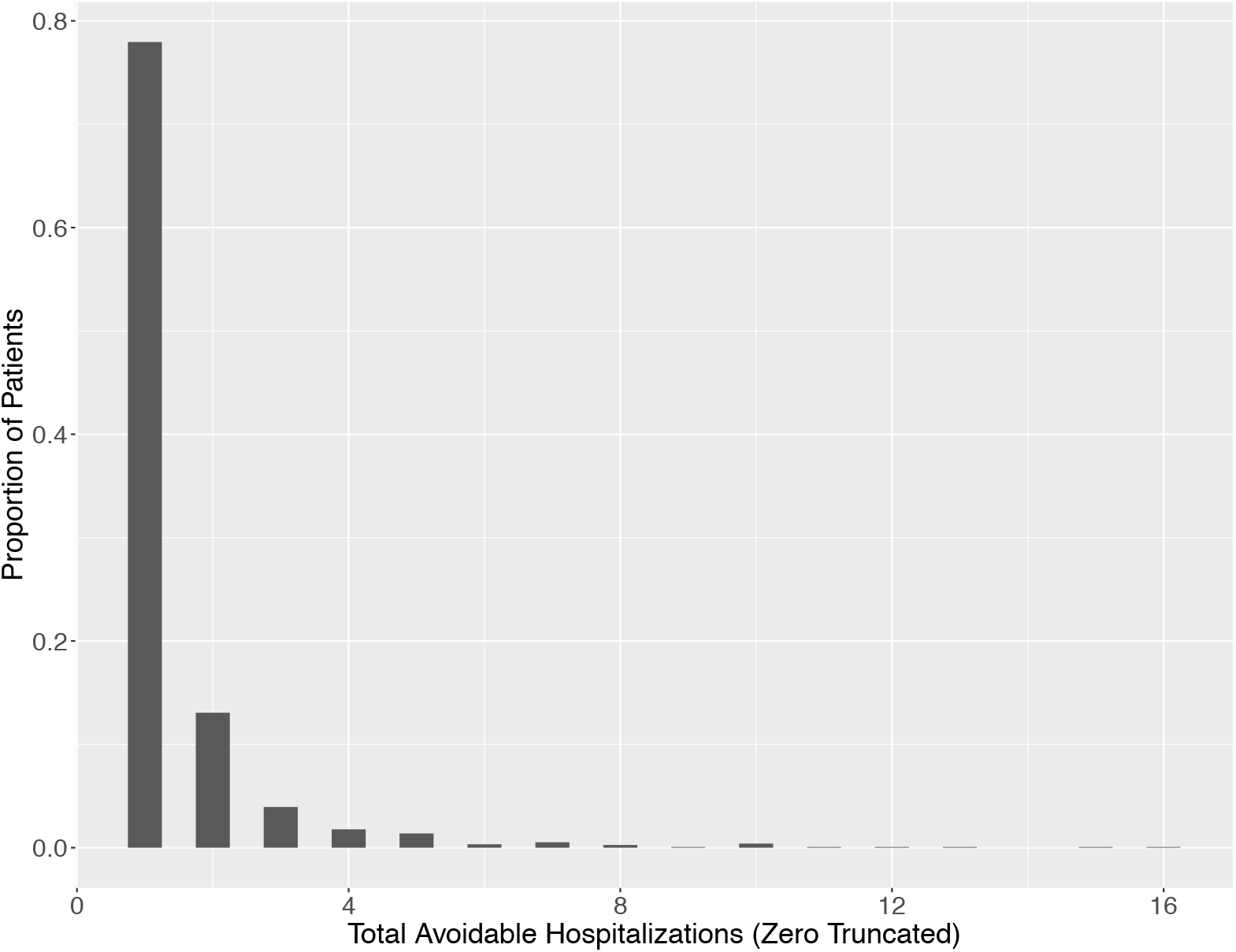
Distribution of total number of avoidable hospitalizations (AHs) for patients with at least one AH in CHOP data sample.

### Analyzing risk factors associated with pediatric avoidable hospitalization

About one-third of pediatric healthcare costs are associated with hospital admissions, the majority of which are unplanned [23]. Unplanned hospitalizations associated with a diagnosis treatable at the primary care level are considered avoidable [24]. By studying which risk factors are most strongly associated with avoidable hospitalizations (AHs), hospital systems can identify patient subpopulations for which primary care should be improved, ideally leading to an overall reduction in hospital costs or admissions [25]. Because pediatric avoidable hospitalization is uncommon, integrating data across hospital systems can lead to more robust inference, increasing power to detect differences in rates of AH among patients. Further, the rarity of pediatric AH makes analyses studying this outcome susceptible to zero-inflation, making this application a suitable use case for hurdle regression.

In this analysis, we applied ODAH to study risk factors associated with pediatric AH using data from the Children’s Hospital of Philadelphia (CHOP) health system. The CHOP system provides care to about 400,000 children per year and includes a large, multi-state outpatient network, as well as one of the largest inpatient facilities for pediatric patients residing in the greater Philadelphia region. Data for this study were extracted from the CHOP EHR system for outpatient, emergency department, and inpatient visits for patients with at least two primary care facility visits from January 2009 to December 2017.

To mimic a scenario in which different sites do not have access to patient-level information at other sites, we assigned patients to the primary care site they attended most often during the study period and carried out analysis as if patient-level information could not be shared across primary care sites. In total, patients were assigned to 27 different primary care sites; we selected six of these sites to illustrate our method, made up of 70,818 patients (Table 2). The largest site of these six, Site 4, was chosen to be the lead site.

**Table 2.** Summary statistics describing patient population across six CHOP primary care sites.

|  | Site 1<br>(n=5456) | Site 2<br>(n=9111) | Site 3<br>(n=7893) | Site 4<br>(n=27288) | Site 5<br>(n=7996) | Site 6<br>(n=13074) | Total<br>(n=70818) |
| --- | --- | --- | --- | --- | --- | --- | --- |
| **Gender** |  |  |  |  |  |  |  |
| Female | 2589<br>(47.5%) | 4427<br>(48.6%) | 3862<br>(48.9%) | 13458<br>(49.3%) | 4013<br>(50.2%) | 6494<br>(49.7%) | 34843<br>(49.2%) |
| Male | 2867<br>(52.5%) | 4684<br>(51.4%) | 4031<br>(51.1%) | 13830<br>(50.7%) | 3983<br>(49.8%) | 6580<br>(50.3%) | 35975<br>(50.8%) |
| **Caucasian Race** |  |  |  |  |  |  |  |
| Caucasian | 3476<br>(63.7%) | 5508<br>(60.5%) | 4783<br>(60.6%) | 15747<br>(57.7%) | 4649<br>(58.1%) | 9158<br>(70.0%) | 43321<br>(61.2%) |
| Other | 1980<br>(36.3%) | 3603<br>(39.5%) | 3110<br>(39.4%) | 11541<br>(42.3%) | 3347<br>(41.9%) | 3916<br>(30.0%) | 27497<br>(38.8%) |
| **Mean Age (across visits) (years)** |  |  |  |  |  |  |  |
| Mean (SD) | 8.02 (5.48) | 7.95 (5.58) | 7.77 (5.50) | 7.54 (5.60) | 7.60 (5.57) | 7.04 (5.37) | 7.57 (5.54) |
| Median [Min, Max] | 7.87<br>[0.0216,<br>18.0] | 7.67<br>[0.0181,<br>18.0] | 7.44<br>[0.0376,<br>17.9] | 6.79<br>[0.0158,<br>17.9] | 7.02<br>[0.0170,<br>17.9] | 6.10<br>[0.0202,<br>17.9] | 6.97<br>[0.0158,<br>18.0] |
| **Insurance Provider** |  |  |  |  |  |  |  |
| Public | 1997<br>(36.6%) | 3410<br>(37.4%) | 2339<br>(29.6%) | 9545<br>(35.0%) | 2477<br>(31.0%) | 3438<br>(26.3%) | 23206<br>(32.8%) |
| Private/Self-Pay | 3459<br>(63.4%) | 5701<br>(62.6%) | 5554<br>(70.4%) | 17743<br>(65.0%) | 5519<br>(69.0%) | 9636<br>(73.7%) | 47612<br>(67.2%) |
| **PC Visits per Year** |  |  |  |  |  |  |  |
| Mean (SD) | 5.19 (5.14) | 5.00 (4.75) | 4.84 (4.35) | 4.51 (4.59) | 5.34 (4.86) | 5.17 (4.85) | 4.88 (4.72) |
| Median [Min, Max] | 3.52 [0.243,<br>65.3] | 3.68 [0.276,<br>85.3] | 3.50 [0.276,<br>70.8] | 3.16 [0.238,<br>97.5] | 3.95 [0.233,<br>73.2] | 3.83 [0.253,<br>85.3] | 3.50 [0.233,<br>97.5] |
| **Hospitalization Status** |  |  |  |  |  |  |  |
| At least one avoidable hospitalization (AH) | 71 (1.3%) | 70 (0.8%) | 33 (0.4%) | 878 (3.2%) | 76 (1.0%) | 396 (3.0%) | 1524 (2.2%) |
| No AHs | 5385<br>(98.7%) | 9041<br>(99.2%) | 7860<br>(99.6%) | 26410<br>(96.8%) | 7920<br>(99.0%) | 12678<br>(97.0%) | 69294<br>(97.8%) |
| **Total AHs (for those with at least one AH)** |  |  |  |  |  |  |  |
| Mean (SD) | 1.38 (1.19) | 1.51 (1.82) | 1.48 (1.64) | 1.46 (1.16) | 1.46 (0.901) | 1.47 (1.58) | 1.47 (1.31) |
| Median [Min, Max] | 1.00 [1.00,<br>10.0] | 1.00 [1.00,<br>15.0] | 1.00 [1.00,<br>10.0] | 1.00 [1.00,<br>10.0] | 1.00 [1.00,<br>5.00] | 1.00 [1.00,<br>16.0] | 1.00 [1.00,<br>16.0] |
| **Follow-up Time** |  |  |  |  |  |  |  |
| Mean (SD) | 3.43 (2.05) | 4.67 (2.76) | 4.74 (2.75) | 4.72 (2.72) | 4.92 (2.77) | 4.75 (2.74) | 4.64 (2.72) |
| Median [Min, Max] | 3.25<br>[0.0766,<br>8.74] | 4.58<br>[0.0766,<br>8.74] | 4.83<br>[0.0766,<br>8.74] | 4.74<br>[0.0766,<br>8.74] | 5.08<br>[0.0766,<br>8.74] | 4.74<br>[0.0766,<br>8.74] | 4.58<br>[0.0766,<br>8.74] |

To evaluate ODAH, we modeled total number of AHs given a collection of EHR variables: gender, race (Caucasian or other), mean age (across all visits), primary care visits per year, and insurance type (public or private). While the majority of patients who experience an AH in these data only experience one, 22% experience more than one, suggesting an advantage of using Poisson regression over logistic regression alone to explicitly model the counts (Figure 3). This, combined with substantial zero-inflation, makes Poisson-Logit hurdle regression appropriate for modeling these data. The logistic component of the hurdle model will model probability of a patient experiencing at least one AH, while the zero-truncated Poisson component will model the total number of hospitalizations for a patient given that they experience at least one.

As in our simulations, we used an identical set of covariates for both hurdle model components and evaluated method performance by calculating relative bias to the pooled estimate for lead site analysis, meta-analysis, and ODAH. To estimate the variance of ODAH parameter estimates, we used the inverse of the Hessian matrix produced when optimizing the surrogate log likelihood function of each hurdle model component.

Figure 4 depicts our AH analysis results. Regression coefficient estimates for each covariate in the fitted hurdle model are shown along with their corresponding 95% confidence interval.

**Figure 4.**
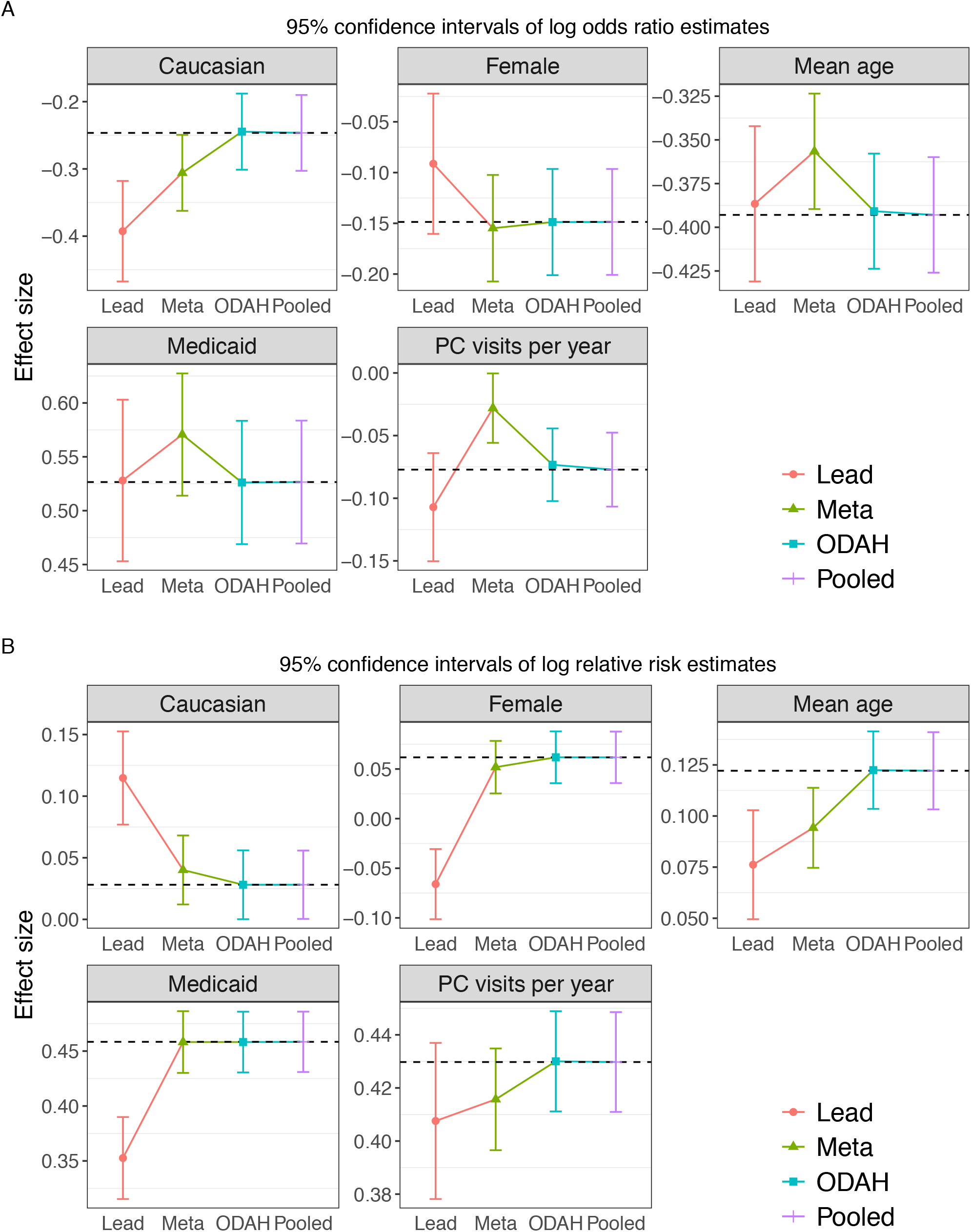
Plots depicting results from CHOP avoidable hospitalization analysis. Log odds ratio (A) and log relative risk (B) estimates (along with corresponding 95% confidence intervals) for each covariate in the fitted hurdle model. Dashed horizontal line represents pooled estimate, our gold standard for comparing methods.

Log odds ratio estimates (estimated by the logistic component of the hurdle model) when using ODAH were close to the pooled estimates, with relative bias ranging from 0.08% (insurance covariate) to 5.02% (primary care visits per year covariate). Meta-analysis estimates were more biased, with relative bias ranging from 4.15% (gender covariate) to 63.6% (primary care visits per year covariate). Log relative risk estimates (estimated by the zero-truncated Poisson component of the hurdle model) were nearly identical when using ODAH and pooled analysis. Meta-analysis performed similarly to ODAH across all coefficients, but ODAH always achieved the smaller relative bias to pooled estimates. ODAH relative bias was < 0.50% for all covariates, while meta-analysis relative bias ranged from 5.89% (PC visits per year) to 11.7% (race).

### Modeling serious adverse event frequency in colorectal cancer patients

Our second analysis studied a population of patients with colorectal cancer (CRC) who use FOLFIRI, an FDA-approved standard of care first line chemotherapy treatment in patients with metastatic CRC, as their CRC treatment. We focused on assessing drug safety in terms of the frequency of serious adverse events (SAEs). The data analyzed are from the OneFlorida Clinical Research Consortium, containing robust longitudinal and linked patient-level real-world data of around 15 million Floridians, making up over 50% of the Florida population. OneFlorida data includes records from Medicaid & Medicare claims, cancer registry data, vital statistics, and EHRs from its clinical partners. These data are centralized in a HIPAA limited dataset that contains detailed patient and clinical variables, including demographics, encounters, diagnoses, procedures, vitals, medications, and labs, following the PCORnet Common Data Model. The OneFlorida data undergo rigorous quality checks at its data coordinating center, the University of Florida, and a privacy-preserving record linkage process is used to deduplicate records of same patients coming from different health care systems within the network [26]. Figure 5 shows the geographic locations of OneFlorida partners.

**Figure 5.**
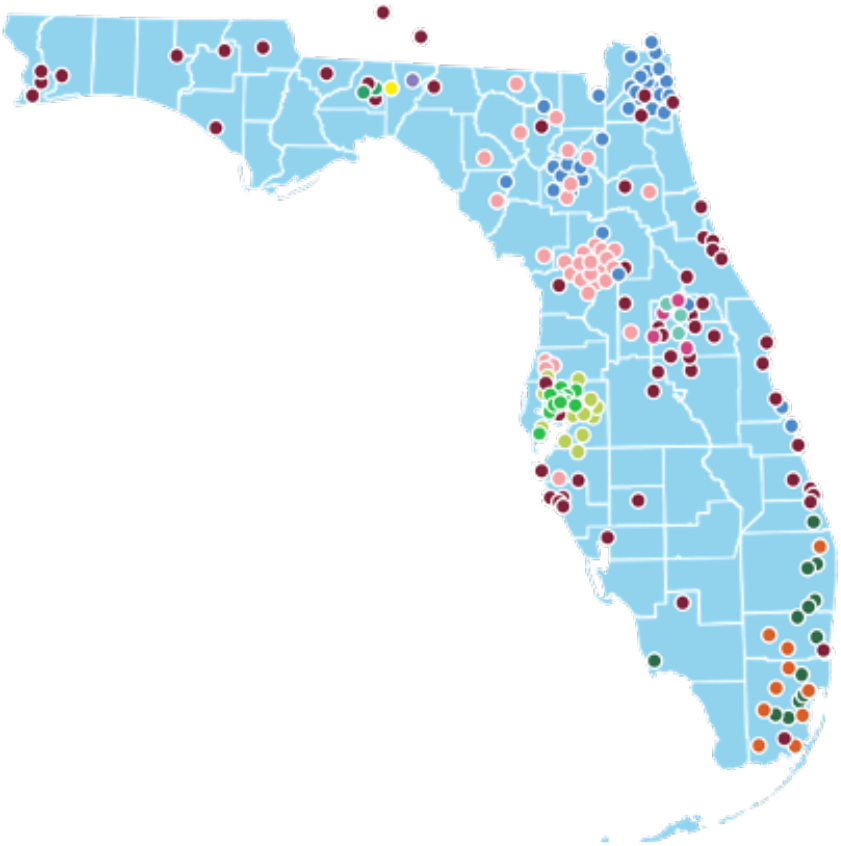
Map detailing locations of OneFlorida clinical partners.

To define an SAE in this analysis, we followed the FDA definition of SAEs and the Common Terminology Criteria for Adverse Events (CTCAE) v 5.0, and the number of SAEs were counted for each patient within 180 days after first FOLFIRI prescription [27]. We removed the chronic conditions that occurred before prescription. A set of covariates and risk factors for all patients were extracted from patients’ medical records for this analysis, including patients’ demographic variables (age, race, Hispanic ethnicity status, and gender) on the day of CRC diagnosis. We also calculated each patient’s Charlson comorbidity index (CCI) using their medical history.

Since OneFlorida data are centralized, we were able to both carry out analysis as if patient-level information could not be shared across clinical sites (as was done in our AH application) as well as fit a hurdle regression model using pooled analysis, which served as the gold standard. In total, our analysis included 660 patients from three clinical sites, with Site 3 being the largest and serving as the lead site (Table 3). To evaluate ODAH using these data, we modeled SAE frequency given the extracted clinical information noted above for each patient. We evaluated method performance as we did for our simulations and AH analysis, again using the same set of covariates in each component of the hurdle model.

**Table 3.** Summary statistics describing patient population across three OneFlorida clinical sites.

|  | Site 1<br>(n=48) | Site 2<br>(n=226) | Site 3<br>(n=386) | Total<br>(n=660) |
| --- | --- | --- | --- | --- |
| **Gender** |  |  |  |  |
| Female | 22 (45.8%) | 90 (39.8%) | 178 (46.1%) | 290 (43.9%) |
| Male | 26 (54.2%) | 136 (60.2%) | 208 (53.9%) | 370 (56.1%) |
| **Caucasian Race** |  |  |  |  |
| Caucasian | 25 (52.1%) | 165 (73.0%) | 302 (78.2%) | 492 (74.5%) |
| Other | 23 (47.9%) | 61 (27.0%) | 84 (21.8%) | 168 (25.5%) |
| **Age (years)** |  |  |  |  |
| Mean (SD) | 51.8 (9.55) | 56.2 (11.9) | 57.2 (11.9) | 56.5 (11.8) |
| **Hispanic** |  |  |  |  |
| Yes | 12 (25.0%) | 9 (4.0%) | 226 (58.5%) | 247 (37.4%) |
| No | 36 (75.0%) | 217 (96.0%) | 160 (41.5%) | 413 (62.6%) |
| **Charlson Comorbidity Index (CCI)** |  |  |  |  |
| Mean (SD) | 5.23 (0.52) | 5.27 (0.75) | 5.24 (0.87) | 5.25 (0.81) |
| **Serious Adverse Events (SAEs)** |  |  |  |  |
| Mean (SD) | 1.81 (1.71) | 2.11 (2.19) | 1.47 (1.72) | 1.72 (1.91) |
| Patients with 0 SAEs | 12 (25%) | 53 (23.5%) | 151 (39.1%) | 216 (32.7%) |
| Zero-Truncated Mean (SD) | 2.42 (1.56) | 2.75 (2.11) | 2.42 (1.61) | 2.55 (1.82) |

Results from using ODAH to model SAE frequency in colorectal cancer patients using data from OneFlorida are shown in Figure 6, displayed similarly to the CHOP AH results. In this application, we again see our method exhibiting low bias relative to pooled estimation. For four of the five log odds ratios estimated in the logistic component of the hurdle model, relative biases produced by ODAH were less than 7%. The lone exception, the gender coefficient, reflected greater relative bias due to its near-zero effect size (reflecting an odds ratio of 1).

**Figure 6.**
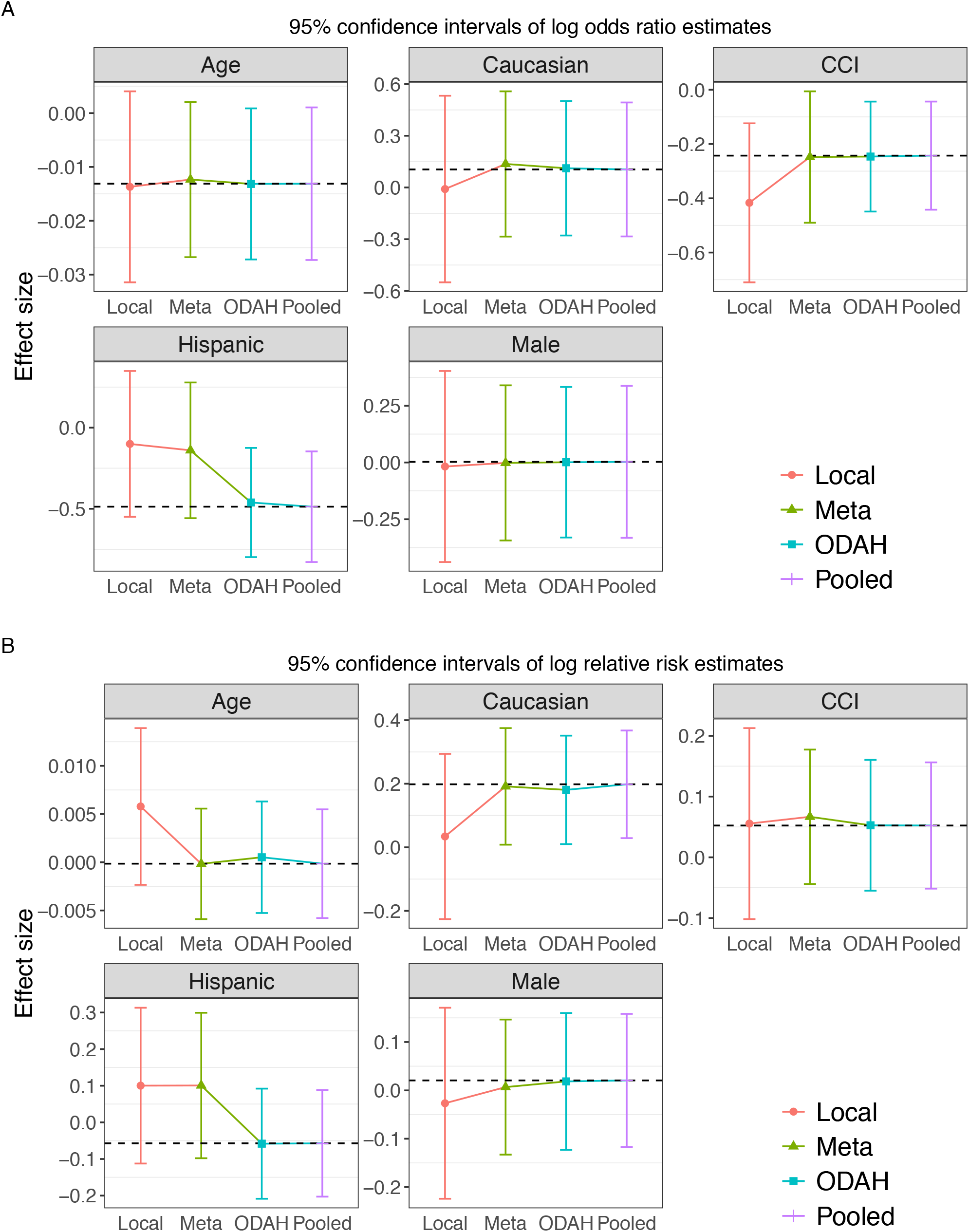
Plots depicting results from OneFlorida serious adverse event application. Log odds ratio (A) and log relative risk (B) estimates (along with corresponding 95% confidence intervals) for each covariate in the fitted hurdle model. Dashed horizontal line represents pooled estimate, our gold standard for comparing methods.

Similar results were observed in the zero-truncated Poisson component, with relative biases to the pooled estimates less than 10% for four of the five estimated log relative risks. The age coefficient had higher relative bias, again due to negligible effect size. In both components, meta-analysis tended to do poorer relative to pooled estimation. The largest difference in estimation can be seen in the coefficients reflecting association of SAE frequency with Hispanic ethnicity, where relative bias was 71% in the logistic component and 276% in the zero-truncated Poisson component (compared to 5.3% and 1.8% for ODAH, respectively).

## Discussion

We introduced a non-iterative, privacy-preserving algorithm for performing distributed hurdle regression with zero-inflated count outcomes. As demonstrated by simulations and two real-world EHR applications, our method consistently produced parameter estimates comparable to and sometimes better than those produced by meta-analysis. Our method’s utility is especially evident in settings featuring a count outcome with marked zero-inflation and very low event rate, as we demonstrated the tendency of only meta-analysis to produce biased estimates under these circumstances in both simulations and real-data analysis. We also showed the benefit of using ODAH over meta-analysis in settings where use of individual site estimates alone may not result in accurate population-level estimation. In the analysis modeling SAE frequency, bias relative to pooled estimation for meta-analysis was greatest for the Hispanic ethnicity coefficient. The proportion of Hispanic patients at each site in this analysis was 4%, 25%, and 58.5%, potentially resulting in highly varied individual-site estimates for this coefficient and resulting in biased meta-analysis estimates. In addition to site-level estimates, our method incorporates aggregate information in the form of gradients to better approximate the complete data likelihood and result in lower bias relative to pooled estimates.

There are several advantages to using our method for performing privacy-preserving data analysis. By using distributed regression, our approach is well-suited for multi-site studies which are ongoing. The surrogate likelihood method takes advantage of patient-level data still being accessible by collaborating sites, allowing collaborators to engage in limited inter-site communication to produce less biased results than would be obtained via meta-analysis, which is best suited for studies already completed. Further, most existing distributed regression techniques require iterative communication among sites to produce accurate estimates. ODAH requires two rounds of non-iterative communication between the local site and all other sites before surrogate likelihood functions can be maximized to obtain accurate, precise parameter estimates. This is particularly advantageous in big data settings, where iterative procedures have a high computational burden in terms of memory and processing time. Further, due to the separability of hurdle model components, each component’s likelihood function can be maximized independently, reducing computational complexity.

Our simulation results suggest that lead site size relative to total population size does not have a discernable effect on any method performance outside of analysis only using data at the lead site. However, since the surrogate log likelihood function only uses individual-level data stored in the lead site, we recommend that the lead site is as large as possible; this helps to ensure the surrogate likelihood is a close approximation to the complete data likelihood.

In terms of limitations of our method, one is that it assumes relative homogeneity among the data to be analyzed. This is an implication of the surrogate likelihood construction, which approximates the complete data log likelihood in part by using a sample-size-weighted sum of gradients from each collaborating site. This implicitly assumes that study data are independent and identically distributed across all sites, which may not hold in some real-world settings. As evidenced by Figure 7, geographical heterogeneity among the patient population can occur in the covariates, with some locations having substantially different demographic makeups than others. We recommend those who implement ODAH ensure patient demographics are largely similar across institutions, or alternatively perform subgroup analysis for relatively homogeneous subsets of institutions. Another limitation is that the zero-truncated Poisson component of our method does not currently account for overdispersion in the outcome; in both of our real data applications, we did not find strong evidence of overdispersion. Finally, there were discrepancies when comparing simulation and data analysis results in terms of bias in the hurdle model’s logistic component estimates. We suspect this is due to simulated data not fully capturing the true distribution of the real data, particularly in terms of covariate imbalance. For example, 52% of patients in the CHOP data that had at least one AH used public insurance, compared to 32% of patients who did not have an AH. We seek to address these limitations in future work.

**Figure 7.**
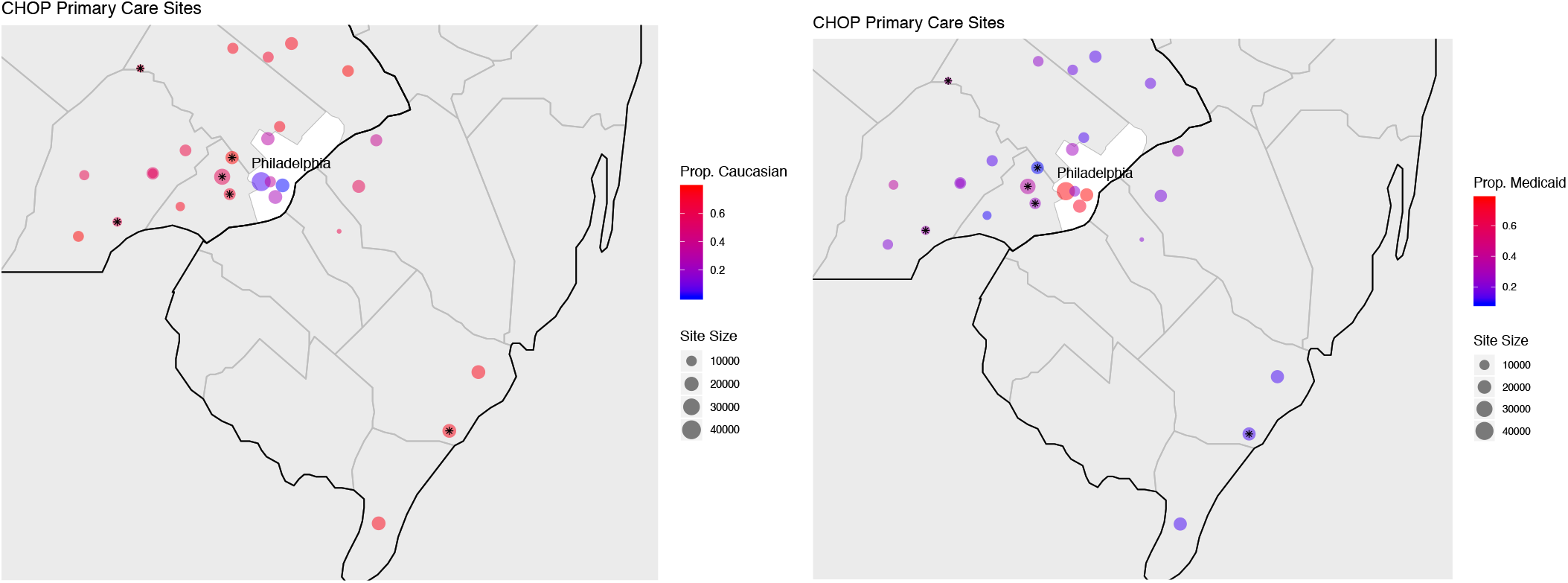
Geographical map of 27 CHOP primary care sites across greater Philadelphia region. In the left map, the proportion of patients of Caucasian race are depicted for each site. In the right map, the proportion of patients using public insurance (Medicaid) at each site is depicted. The size of each site on each map is proportional to the number of patients at the given site. Stars indicate sites used in our data analysis.

Our group continues to construct methods for performing non-iterative, privacy-preserving distributed inference, ideal for use within CRNs which seek to collaborate on analyses without sharing patient-level data. We seek to create distributed methods for the types of outcomes most common in healthcare, so far producing methods for modeling binary [17], time-to-event [20], and now zero-inflated count outcomes. While additional methods are being developed and implemented, we believe ODAH is worthy of consideration when one seeks to perform distributed regression on zero-inflated count outcome data.

## Conclusion

We proposed an accurate, efficient, privacy-preserving algorithm (ODAH) for performing distributed hurdle regression for settings featuring a zero-inflated count outcome. By only requiring patient-level data from one site, we limit between-site communication to sharing only aggregate statistics, preserving patient privacy. In an extensive simulation study and two real-world data analyses, ODAH exhibited higher estimation accuracy than meta-analysis, most notably in the context of rare events. We believe ODAH can be a useful method for analyzing zero-inflated count outcomes in a clinical research network where patient-level data cannot be shared.

## Methods

### Poisson-Logit Hurdle Model

A hurdle model is a two-part model which specifies two separate processes, one for generating zero values and another for generating values given that they are non-zero [28]. The hurdle model is useful for modeling a count outcome with excess zeros, modeling the zero and positive counts independently. Figure 8 depicts a typical zero-inflated distribution of counts, as well as a schematic overview detailing the sequential nature of the Poisson-Logit hurdle model.

**Figure 8.**
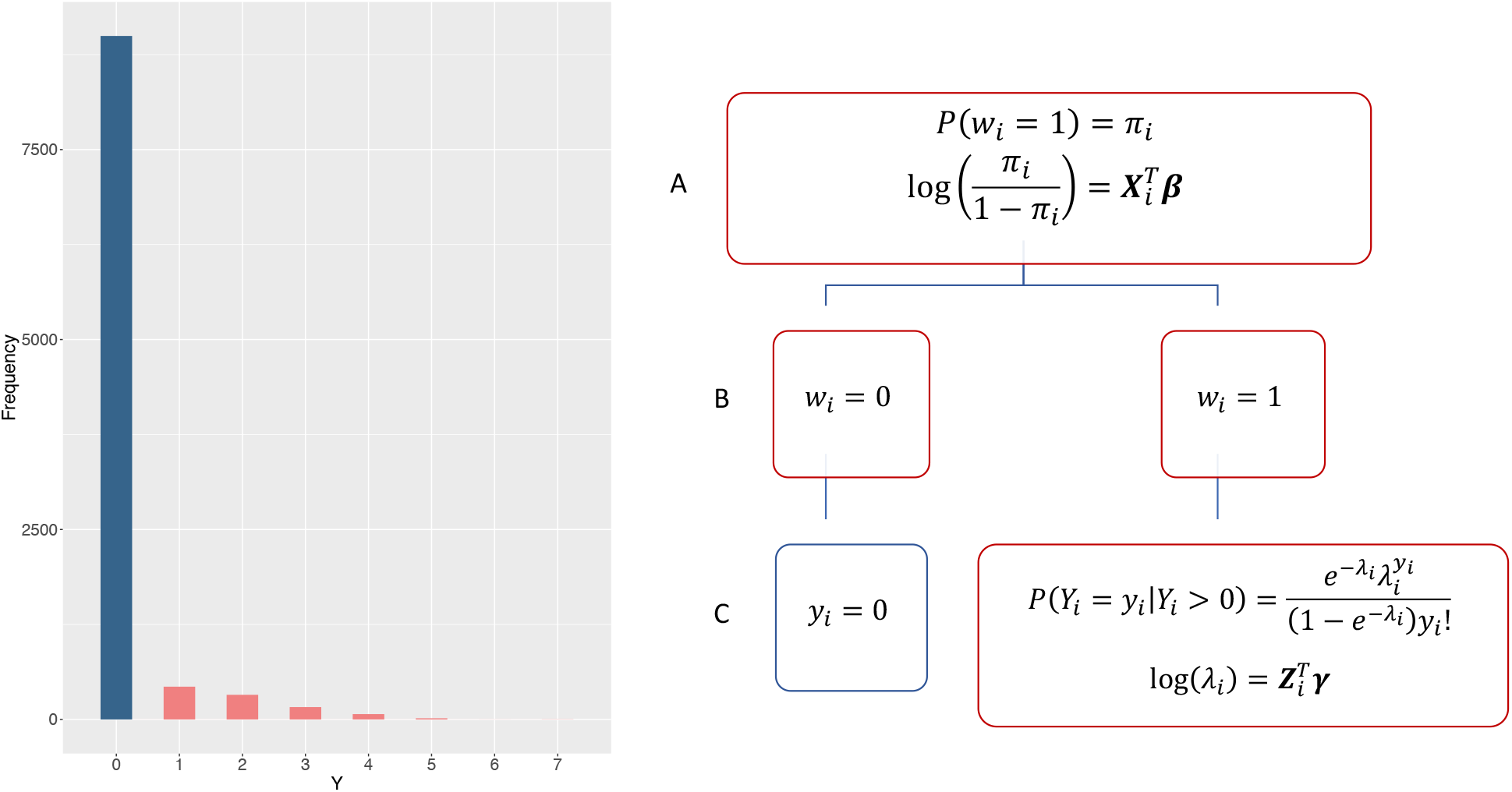
On the left, a histogram displaying counts generated with a Poisson-Logit hurdle distribution with 10% prevalence and a zero-truncated event rate of *λ* = 1.5. On the right, a hierarchical diagram visualizing the data generation process in a Poisson-Logit hurdle framework. Independent realizations *w*_*i*_ ∈ {0,1} are generated from a Bernoulli process, with underlying probability *π*_*i*_ modeled using a logit link. Realizations where *w*_*i*_ = 0 are zero counts (*y*_*i*_ = 0), while realizations where *w*_*i*_ = 1 are positive counts (*y*_*i*_ ∈ {1,2, …}). The positive counts are generated by a zero-truncated Poisson distribution, with underlying event rate *λ*_*i*_ modeled using a log link.

In this paper, we invoke the hurdle model to model zero-inflated count outcomes common in healthcare data. To derive our hurdle model, we consider the two processes making up the model independently. First, we model the proportion of zero counts with a Bernoulli process using a logit link. Let *w*_*i*_, *w*,…, *w*_*n*_ ∈ {0,1} be independent realizations of a binary response variable *W*, such that *P*(*w*_*i*_ = 1) = *π*_*i*_ and *P*(*w*_*i*_ = 0) = 1 − *π*_*i*_. The logistic model of the probability *π*_*i*_ is modeled as a linear combination of explanatory variables ***X*** and regression coefficients ***β***:

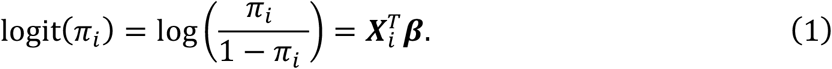

Next, positive counts are modeled using a zero-truncated Poisson model. Let *y*_1_, *y*_2_,…, *y*_*n*_ ∈ {0,1,2, …} be independent realizations of a count variable *Y*. Assume *P*(*Y*_*i*_ = 0) = *P*(*w*_*i*_ = 0) = 1 − *π*_*i*_, and *P*(*Y*_*i*_ > 0) = *P*(*w*_*i*_ = 1) = *π*_*i*_. Thus, *π*_*i*_ can interpreted as the probability that the “hurdle is crossed”, resulting in a non-zero count. In the context of zero-inflated counts, we assume *P*(*y*_*i*_ = 0) is much greater than *P*(*y*_*i*_ > 0).

For observations where the realization from the logistic model is 1, positive counts follow a zero-truncated Poisson distribution such that 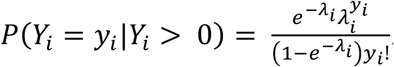. Thus, we can write the mixture probability mass function of the Poisson hurdle model as

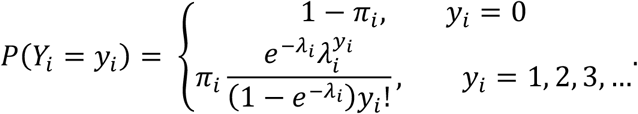

Modeling the rate parameter *λ*_*i*_ using a log link, we can express the log of *λ*_*i*_ as a linear combination of explanatory variables ***Z*** and regression coefficients ***γ***:

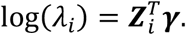

We write the log-likelihood of the Poisson hurdle model as *L*(***β, γ***) = *L*_*i*_(***β***) + *L*_2_(***γ***), with

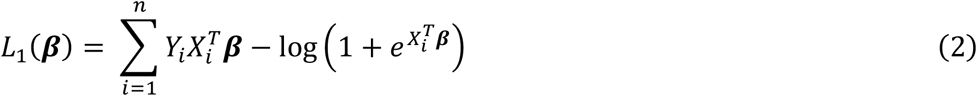

and

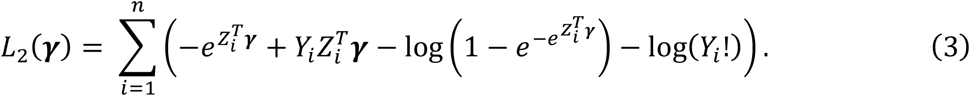

Note that this factors into two components such that **β** and **γ** are separable; the Hessian matrix is block diagonal, so **β** and **γ** are information orthogonal. Thus, there will not be any loss of information in estimating each set of parameters separately. This property is useful in the context of distributed regression, reducing computational complexity.

While less common than traditional regression models for count data, hurdle models have been used successfully in various health contexts with substantial zero inflation. For instance, Negative Binomial – Logit hurdle models were utilized to estimate risk of vaccine adverse events for clinical trial participants, as well as to estimate cigarette and marijuana use among youth e-cigarette users [29-30]. Hurdle regression has also been used in other specialized contexts, such as in estimating spatiotemporal patterns of emergency department use and quantifying association between preventive dental behaviors and caries prevalence [31-32]. Contrary to zero-inflated Poisson or Negative Binomial regression models, hurdle models have only one source of zero counts, indicating that all individuals in the study sample are at risk of the outcome. This offers an interpretation of estimated model coefficients that is more appropriate in many clinical settings.

### Distributed Hurdle Regression: ODAH

Suppose we have clinical data stored in *K* sites, where the *j*^th^ site has a sample size *n*_*j*_ and the total sample size across sites is 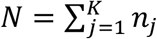. Let *Y*_*ij*_ and *X*_*ij*_ denote the count outcome and covariate vector for subject *i* in site *j*, respectively. We can write the log likelihood functions for the combined data as

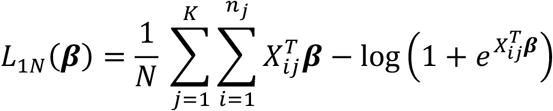

and

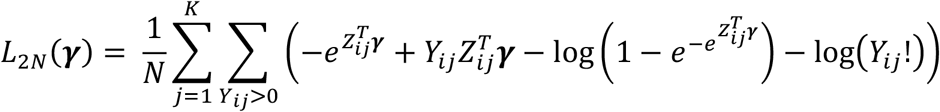

In the CRN context, we assume that we do not have access to the combined data. We only have access to data at one of the *K* sites (the *lead* site, with site index *j* = 1), as well as aggregate information from the other sites (the *collaborating* sites). Using methods developed by Jordan et al. and later adapted to the clinical data setting by Duan et al., we construct a *surrogate log likelihood function*, which approximates the complete data log likelihood using patient-level data from the lead site and aggregate information from the collaborating sites [17, 20-21]. The aggregate information used in our work is the set of first- and second-order gradients of the log likelihood function at the *K* – 1 collaborating sites. The surrogate log likelihood function for each component of the hurdle model can be expressed as

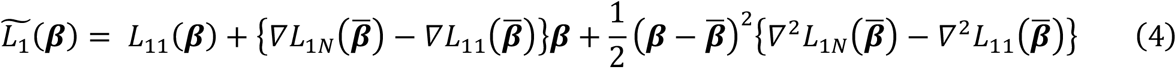

and

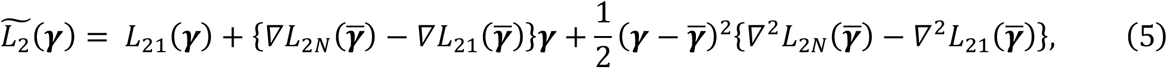

where 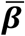 and 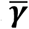 are initial estimates for the algorithm. Here, *L*_11_(***β***) and *L*_21_(***γ***) are log-likelihoods computed using patient-level data at the lead site for the logistic and zero-truncated components, respectively. The terms

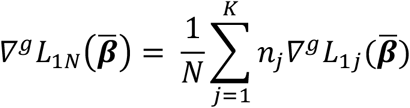

and

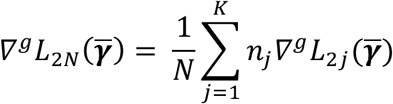

are weighted averages of first-order (*g* = 1) or second-order (*g* = 2) gradients at each site, and 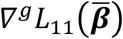 and 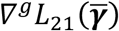 are first-order or second-order gradients calculated at the lead site for the logistic and zero-truncated Poisson components of the hurdle model, respectively, evaluated at 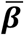 and 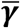. Explicit formulations of the first- and second-order gradients for each component of the hurdle model are available in the Supplement (Equations S.1 – S.4). The ODAH estimators are then defined as

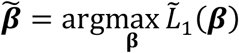

and

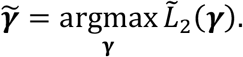

Well-chosen 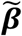 and 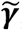 will increase the accuracy of 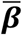 and 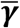, respectively. In this work, 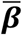 and 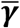 are estimates computed from performing a fixed-effects meta-analysis using all *K* sites, or inverse-variance weighted sums of estimates from the *K* studies, i.e. for 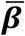 (with 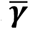 similar),

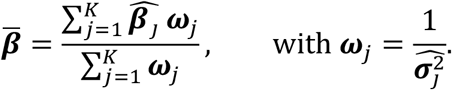

This requires each site to send point and variance estimates to the lead site to initiate the algorithm. Alternatively, one could use lead site maximum likelihood estimates 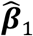 and 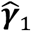 obtained via fitting the hurdle model of interest at the lead site. This has been shown to perform well when the lead site is largely representative of the entire multi-site sample and eliminates one round of communication among sites relative to using the meta-analytic initial estimate [17].

When using a meta-analysis estimate to initiate ODAH, two non-iterative rounds of communication are necessary for transferring information across sites; thus, our approach is considered a *one-shot* approach for performing distributed regression. ODAH requires each collaborating site to first fit the hurdle model of interest using its own data before sending parameter point and variance estimates to the lead site. A user at the local site can then initiate ODAH by, following its own hurdle model fitting, computing initial estimates via meta-analysis before sending these estimates to the collaborating sites for computing gradients. These gradients are then sent to the lead site to construct the surrogate log likelihood function. Using only gradients and patient-level data from the lead site, we obtain parameter estimates calculated from maximizing each surrogate likelihood function with respect to the parameter of interest. The ODAH algorithm is outlined in detail below.

## Supporting information

Supplemental Equations and Simulation Results

## Data Availability

Children's Hospital of Philadelphia: The data are not available for deposit into a repository. CHOP can make aggregate results available on request.
OneFlorida: OneFlorida data can be requested at https://onefloridaconsortium.org/front-door/; Since OneFlorida data is a HIPAA limited data set, a data use agreement needs to be established with the OneFlorida network.

## Algorithm

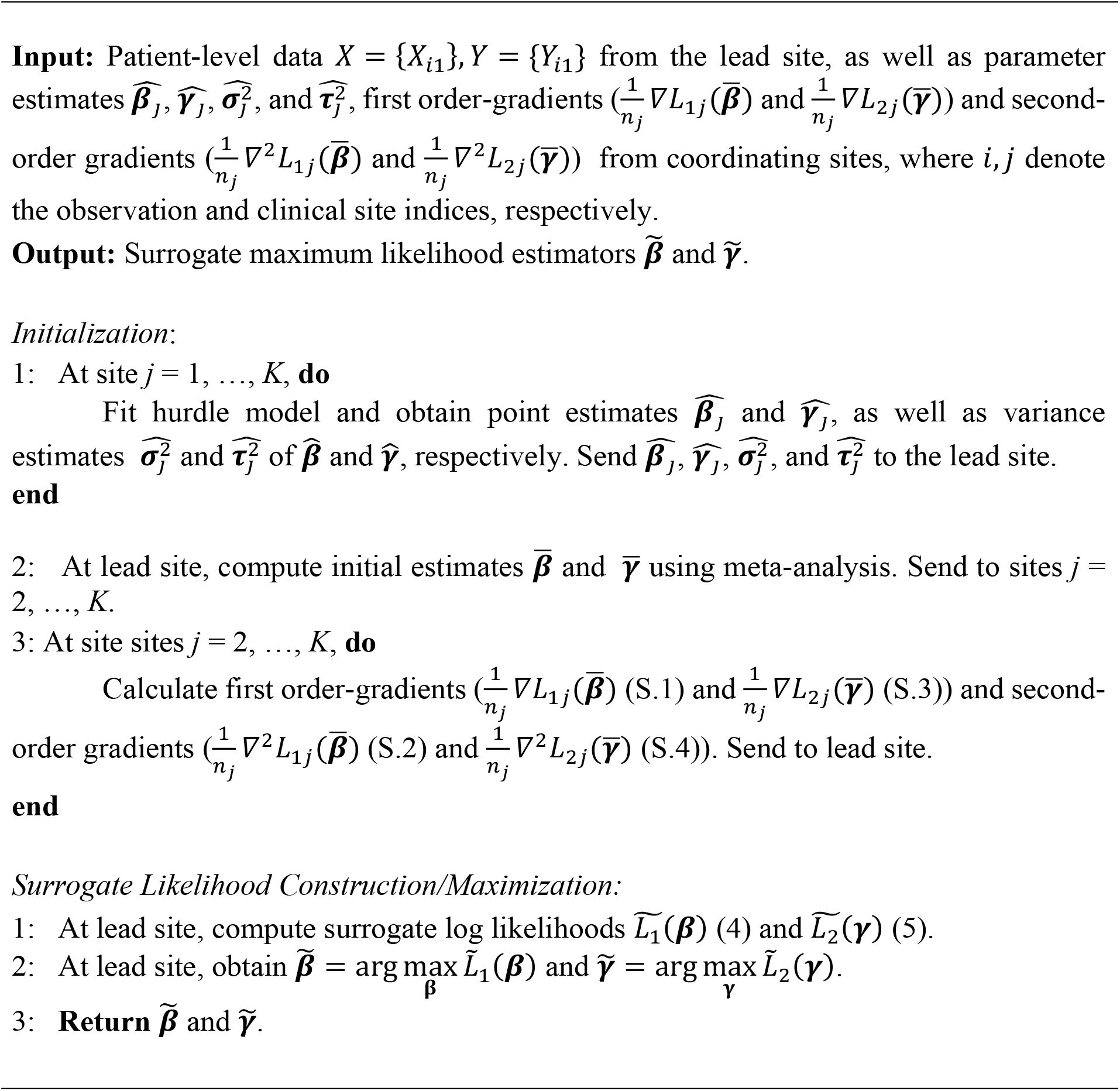

