## Supplemental Equations and Simulation Results for "An Efficient and Accurate Distributed Learning Algorithm for Modeling Multi-Site Zero-Inflated Count Outcomes"

Supplementary Material

S.1) First-and second-order gradient formulations

**Logistic component:**

$$\nabla l_1(\boldsymbol{\beta}) = \frac{\partial}{\partial \beta} l_1(\boldsymbol{\beta}) = \sum_{i=1}^{n} \boldsymbol{X}_i - \sum_{i=1}^{n} \frac{\exp(\boldsymbol{X}_i^t \boldsymbol{\beta})}{1 + \exp(\boldsymbol{X}_i^T \boldsymbol{\beta})} \boldsymbol{X}_i \qquad (\text{S}.1)$$

$$\nabla^2 l_1(\boldsymbol{\beta}) = \frac{\partial^2}{\partial \boldsymbol{\beta} \partial \boldsymbol{\beta}^T} l_1(\boldsymbol{\beta}) = -\sum_{i=1}^{n} \frac{\exp(\boldsymbol{X}_i^T \boldsymbol{\beta})}{(1 + \exp(\boldsymbol{X}_i^T \boldsymbol{\beta}))^2} \boldsymbol{X}_i \boldsymbol{X}_i^T \qquad (\text{S}.2)$$

**Zero-truncated Poisson component:**

$$\nabla l_2(\boldsymbol{\gamma}) = \frac{\partial}{\partial \boldsymbol{\gamma}} l_2(\boldsymbol{\gamma}) = \sum_{Y_i > 0} \left( -\exp(\boldsymbol{Z}_i^T \boldsymbol{\gamma})\, \boldsymbol{Z}_i + y_i \boldsymbol{Z}_i - \frac{\exp(-\exp(\boldsymbol{Z}_i^T \boldsymbol{\gamma}))\exp(\boldsymbol{Z}_i^T \boldsymbol{\gamma})\, \boldsymbol{Z}_i}{1 - \exp(-\exp(\boldsymbol{Z}_i^T \boldsymbol{\gamma}))} \right) \qquad (\text{S}.3)$$

$$\nabla^2 l_2(\boldsymbol{\gamma}) = \frac{\partial^2}{\partial \boldsymbol{\gamma} \partial \boldsymbol{\gamma}^T} l_2(\boldsymbol{\gamma}) = \sum_{Y_i > 0} -\exp\left(\boldsymbol{Z}_i^T \boldsymbol{\gamma}\right) \boldsymbol{Z}_i \boldsymbol{Z}_i^T + \sum_{Y_i > 0} \left( \frac{\exp\left(\boldsymbol{Z}_i^T \boldsymbol{\gamma}\right) \left[\exp\left(\exp\left(\boldsymbol{Z}_i^T \boldsymbol{\gamma}\right) + \boldsymbol{Z}_i^T \boldsymbol{\gamma}\right) - \exp\left(\exp\left(\boldsymbol{Z}_i^T \boldsymbol{\gamma}\right)\right) + 1\right]}{\left[\exp\left(\exp\left(\boldsymbol{Z}_i^T \boldsymbol{\gamma}\right)\right) - 1\right]^2} \boldsymbol{Z}_i \boldsymbol{Z}_i^T \right) \qquad (\text{S}.4)$$

### S.2) Additional simulation results

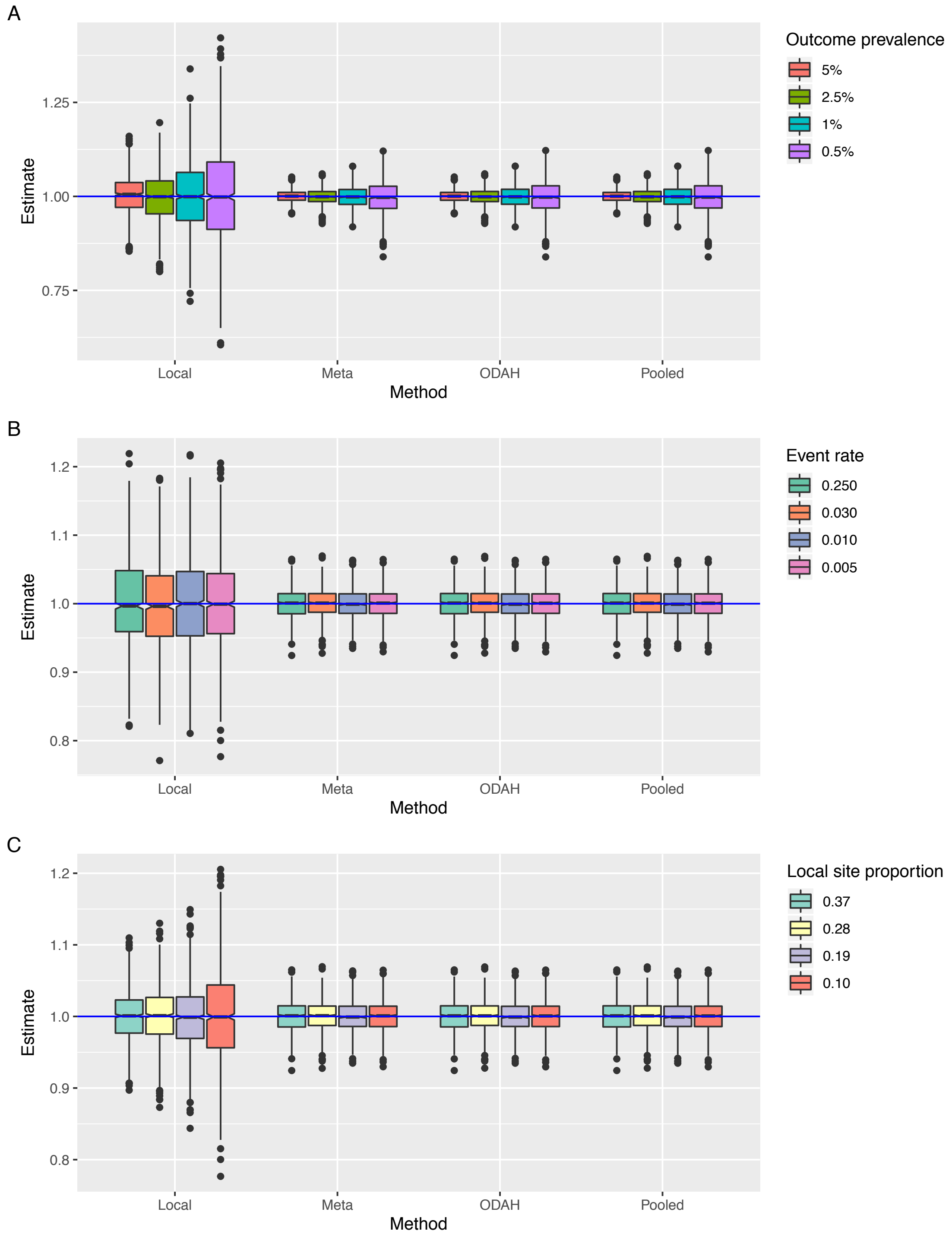


Figure S.2. Simulation results for estimating logistic component covariate $\beta_2$. A) Results for Setting A, fixing $n_{local}$ = 20,000 and $\gamma_0$ = -3.6 ($\lambda = 0.03$) while varying outcome prevalence. B) Results for Setting B, fixing $n_{local}$ = 20,000 and $\beta_0$ = -3.7 (2.5% prevalence) while varying event rate ($\lambda$). C) Results for Setting C, fixing $\beta_0$ = -3.7 (2.5% prevalence) and $\gamma_0 = -3.6$ ($\lambda = 0.03$) while varying proportion of observations in local site. Horizontal blue line represents true value of $\beta_2 = 1$.
